# BASH-GN: A new machine learning derived questionnaire for screening obstructive sleep apnea

**DOI:** 10.1101/2022.02.05.22270403

**Authors:** Jiayan Huo, Stuart F. Quan, Janet Roveda, Ao Li

## Abstract

**Purpose:** This study aims to develop a machine learning based questionnaire (BASH-GN) to classify obstructive sleep apnea (OSA) risk by considering risk factor subtypes.

**Methods:** A total of 4,527 participants that met study inclusion criteria were selected from Sleep Heart Health Study Visit 1 (SHHS 1) database. Another 1,120 records from Wisconsin Sleep Cohort (WSC) served as an independent test data set. Participants with an apnea hypopnea index (AHI) ≥ 15/h were considered as high OSA risk. Potential risk factors were ranked using mutual information between each factor and the AHI, and only the top 50% were selected. We classified the subjects into 2 different groups, low- and high phenotype groups, according to their risk scores. We then developed the BASH-GN, a machine learning based questionnaire that consists of two logistic regression classifiers for the 2 different subtypes of OSA risk prediction.

**Results:** We evaluated the BASH-GN on the SHHS 1 test set (n = 1237) and WSC set (n = 1120) and compared its performance with four commonly used OSA screening questionnaires, the Four-Variable, Epworth Sleepiness Scale, Berlin, and STOP-BANG. The model outperformed these questionnaires on both test sets regarding the area under the receiver operating characteristic (AUROC) and the area under the precision-recall curve (AUPRC). The model achieved AUROC (SHHS 1: 0.78, WSC: 0.76) and AUPRC (SHHS 1: 0.72, WSC: 0.74), respectively. The questionnaire is available at: https://c2ship.org/bash-gn

**Conclusion:** Considering OSA subtypes when evaluating OSA risk can improve the accuracy of OSA screening.

## Introduction

Obstructive sleep apnea (OSA) is one of the most common sleep disorders and has a significant negative impact on health [1]. It is estimated that 25% of American adults are affected by OSA [2]. Patients with OSA suffer from symptoms, such as excessive daytime sleepiness and insomnia, and have a significant comorbidity burden. Studies have found that OSA patients show a high prevalence of cardiovascular diseases [3], diabetes [4], and depression [5].

Despite improved awareness of OSA, 75-80% of the OSA cases remained undiagnosed [6]. In-lab polysomnography (PSG) is considered as the gold standard for OSA diagnosis. It records multiple physiologic signals that are indicators of sleep architecture and quality, respiration, cardiac rhythm, and movement. Although less costly and intrusive, type III and type IV portable monitors, as substitutes for PSG, are commonly used to diagnose OSA at home. However, they still incur cost and require specific expertise to process and interpret [7]. Due to the large number of patients with suspected OSA, evaluating all suspected OSA patients will lead to long waiting times for testing and high costs.

To alleviate the above problem, there has been substantial research into developing screening processes to identify the patients who should be tested further with PSG. Several screening tools utilizing symptom severity and other risk factors have been proposed to identify patients with high OSA risk. The Epworth Sleepiness Scale (ESS) has been used to determine potential sleep disorders for patients based on 8 sleepiness questions [8]. Takegami et al proposed a 4-variable tool to identify sleep disorders severity [9]. The tool calculates the score using gender, body mass index (BMI), snoring, blood pressure, and their corresponding weights. The Berlin questionnaire (BQ) consists of three sections: snoring, daytime fatigue, and hypertension and BMI [10]. If two or more sections are evaluated as positive, the patient is considered high risk for OSA. The STOP-BANG, one of the most widely accepted screening tools for OSA, utilizes 8 questions to evaluate OSA risk [11]. However, studies show that OSA has different clinical subtypes regarding symptoms [12, 13]. Current screening questionnaires do not consider OSA subtypes and classify subjects using the same standard, resulting in some inaccuracy.

In this study, our hypothesis is a better screening performance can be achieved by customizing the screening process by considering different subtypes of OSA. Therefore, we developed and evaluated a machine learning based questionnaire (BASH-GN) that takes OSA subtypes into account to classify OSA risk.

## Method

### Data sources

The Sleep Heart Health Study (SHHS) was a multi-center cohort study to determine the cardiovascular and other sleep disordered breathing consequences [14]. It recorded full overnight at-home PSG and acquired Sleep Health Questionnaires of 6,441 men and women aged 40 years and older between 1995 and 1998 during the first visit, with 5,804 studies available for analysis. We used the SHHS Visit One (SHHS 1) to develop and test the model. The Wisconsin Sleep Cohort (WSC) database was used as an independent test set to evaluate the generalizability of the model. The WSC is an ongoing longitudinal study of causes and consequences of sleep apnea [15] using overnight in-laboratory studies with a baseline sample of 1,500 Wisconsin state employees. A detailed description of the two datasets is available on the National Sleep Research Resources (NSRR) [16] website.

### Data preprocessing

Risk factors associated with OSA were used as the input features of the model. First, we identified potential risk factors through literature search. We secondly excluded risk factors that are not easy-accessible or suitable for questionnaires. The remaining risk factors included gender[17], BMI[18], snoring[19], age[17], stroke[19], neck girth[20], ethnicity[17], daytime sleepiness[21], alcohol[3], diabetes[3], coronary artery diseases[3], craniofacial change[17], genetics[19], cardiac arrhythmias[3], nasal congestion[19], night sweats[20], smoking[19], sleep quality[18], obesity[18], hypothyroidism[3], acromegaly[3], large tonsils[3], menopause[19], and hypertension[22].

In the next step, we excluded a total of 1,681 SHHS 1 subjects due to missing values of risk factors related to OSA or variables that would be used in the Four-Variable, ESS, Berlin, and STOP-BANG questionnaires for comparison. These data appeared to be missing at random. The variables and missingness frequency are provided in Fig. 1. The final SHHS 1 dataset consisted of 4,123 participants. The first visit of the WSC database contained 1,123 participants, of which 3 were excluded due to missing ESS score or diastolic pressure. The 4,123 participants selected from SHHS 1 dataset were randomly split into training and testing sets in a ratio of 7:3. The 1120 subjects from WSC served as the independent test set.

**Fig. 1.**
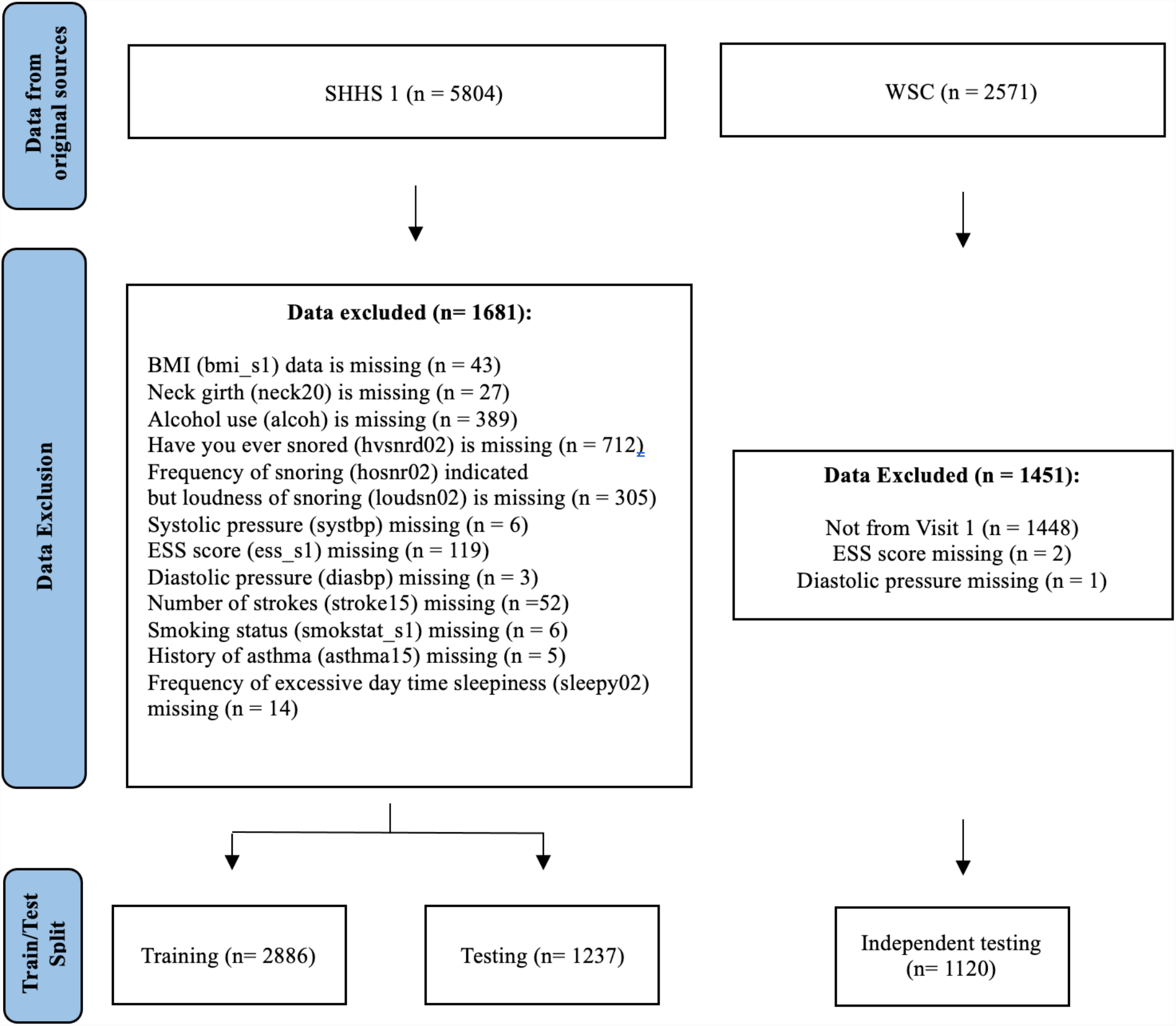
Flow chart of data inclusion in this study. BMI: body mass index; ESS: Epworth Sleepiness Scale

We classified OSA severity according to the apnea hypopnea index (AHI) as previously described [23]. Specifically, AHI with ≥3% oxygen desaturation or arousal was used as the ground truth, based on which OSA severity can be defined as minimal (AHI < 5/h), mild (5/h≤AHI<15/h), moderate (15/h≤AHI<30/h) and severe (AHI≥30/h). To compare to performance of our new model with previous questionnaires, the model made a binary classification in which minimal and mild was marked as low risk with label 0 while moderate and severe was considered as high risk and marked with label 1.

Feature selection subsequently was conducted to reduce the complexity of the model and questionnaire. First, we converted the original AHI to the binary AHI severity label (0 for low risk and 1 for high risk) using a cut-off value of 15/h. After the exclusion process, snoring frequency and snoring loudness may still be missing if the participant answered *No* to snoring history. Therefore, we replaced these missing values with 1 and 0 to denote *Do not snore anymore* and *Do not snore*, respectively. Furthermore, snoring frequency was treated as *Do not snore anymore* if the participant answered *Don’t know*. Finally, all other variables used their original values as recorded in the SHHS 1 dataset. Considering the different data types and distributions of the risk factors, we calculated the normalized mutual information (MI) score between each risk factor and binary AHI severity using the equation described by Ross [24]. The MI measures the amount of information that one random variable contains about the target variable. High MI means a large reduction in the uncertainty of the target variable when the values of a random variable are provided. Zero MI means the two variables are independent. The risk factors then were ranked in descending order by MI score.

Corresponding variables were chosen from WSC to match the selected risk factors for independent testing. It should be noted that WSC separated AHI ≥3% oxygen desaturation with or without arousal into rapid eye movement (REM) and Non-REM stages. We calculated the sum AHI of these two stages as the ground truth and used the same cut-off value, 15 /h, to convert the AHI to the binary label. To compare with the previous questionnaires, we also extracted the variables that are being used in STOP-BANG (snoring loudness, tiredness, observed apnea, high blood pressure, BMI, age, neck girth and gender), ESS and Four-Variable (BMI, gender, systolic/diastolic blood pressure, snoring frequency), Berlin (snoring, sleepiness/fatigue, hypertension, BMI). A detailed utilization of variables in both datasets is described as Supplementary Table S1. Re-coding of these variables for use in the STOP-BANG, ESS, Four-Variable and Berlin questionnaires is described in the supplement. The categorical variable snoring loudness was binary encoded. Genders were relabeled for female as 0 and male as 1. Continuous variables, including age, neck girth, and BMI, were standardized to improve the prediction.

### Model development

#### Phenotype classification

We developed a machine learning model to identify the OSA risk. The model used answers of questionnaire as input to predicted subjects as high- or low-risk for OSA. A minimally symptomatic OSA subtype, as described by Keenan et al. [12] and Kim et al. [13], is challenging to screen using a questionnaire due to the lack of the cardinal symptoms associated with OSA. To enhance the performance of prediction in this population with fewer symptoms or findings related to OSA, we firstly divided the subjects into two groups, a low phenotype group and a high phenotype group, according to their answers in the SHHS 1 questionnaire. Specifically, each question was assigned a score of 1 and then we used the following cut-offs for scoring: gender = male, neck circumference > 40 cm [25], age > 50 years [26], BMI > 35 kg/m^2^ [26], high blood pressure = Yes, snoring louder than talking [27]. A score of 2 or less, determined by the area under the receiver operating characteristic (AUROC) (as shown in Fig. S1), out of a total possible score of 6 was considered as low phenotype while a score of 3 and above was considered as high phenotype in this study. Fig. S2 highlights the phenotypic differences. Then we used two independent sub-models for each group to customize the classification process.

#### Algorithm selection

We used stratified 10-fold cross-validation to explore the best algorithm for each sub-model from 8 candidate algorithms, including logistic regression (LR), support vector classifier (SVC), K-nearest neighbors (KNN), decision tree (DT), extra tree (ET), Ada boost (AB), Gaussian Naïve Bayes (GNB) and random forest (RF). Logistic regression had the best AUROC performance in both subtypes as shown in Fig. S3. Thus, the final selected BASH-GN model employed a scoring threshold of 2 to split the subjects into two subtypes, followed by two independent logistic regression classifiers with L2 regularization for each subtype of OSA risk prediction. Then, we trained the two independent logistic regression classifiers on the whole training set (n=2,886) from SHHS 1. The models were implemented by Python v3.8 with package Scikit-learn v0.24.

#### Model evaluation

We evaluated the BASH-GN model on the holdout test set (n = 1,237) and compared the BASH-GN model with STOP-BANG, ESS, Berlin, and Four-variable questionnaires on the area under the precision-recall curve (AUPRC) and AUROC. Then, we applied the pre-trained model on WSC to test the generalizability of the model. We used the same decision threshold (p = 0.427) in the holdout test set to predict OSA risk for WSC set. Finally, AUPRC and AUROC were calculated based on prediction results. The details of STOP-BANG, ESS, Berlin, and Four-variable questionnaires are described in the Supplement.

### Statistical analysis

We used mean and standard deviation as well as percentages to provide an overall description of the training and test sets. We used t-test and Cohen’s d to calculate p values and effect sizes for continuous variables. Chi-square test and Cohen’s w were employed to calculate p values and effect sizes for categorical variables. We considered that p value < 0.05 and effect size > 0.3 indicated statistical significance in our analysis. The AUROCs were used as the metric to evaluate performance. The AUROC shows the true positive rate (sensitivity) versus the false positive (1-specificity) rate when probability thresholds vary. In cases of imbalanced OSA risk distribution, AUPRC can give a more informative picture of an algorithm’s performance [28] as it focuses on positive cases. The precision-recall curve (PRC) shows the precision versus the recall (sensitivity) rate when probability thresholds vary. Thus, we also report AUPRC with 95% confidence intervals (CI) of the BASH-GN model and the comparison questionnaires on both testing sets. A bootstrapping (n = 1000) was used to estimate the 95% CI for each model/questionnaire metrics. Analyses were performed using Python v3.8 with package Scikit-learn v0.24 and SciPy v1.6.

## Results

Table 1 describes the demographic, anthropometric and clinical characteristics of datasets. The asterisk in Table 1 denotes the significant difference regarding variable distribution between two testing sets. The descriptive characteristics between SHHS 1 testing and WSC showed differences, especially in age, BMI and AHI label. A total of 51.07% in WSC were classified as low-risk of OSA and 54.81% were low-risk in SHHS 1 testing set (p value = 3.23×10^−8^, effective size = 4.98).

**Table 1.**
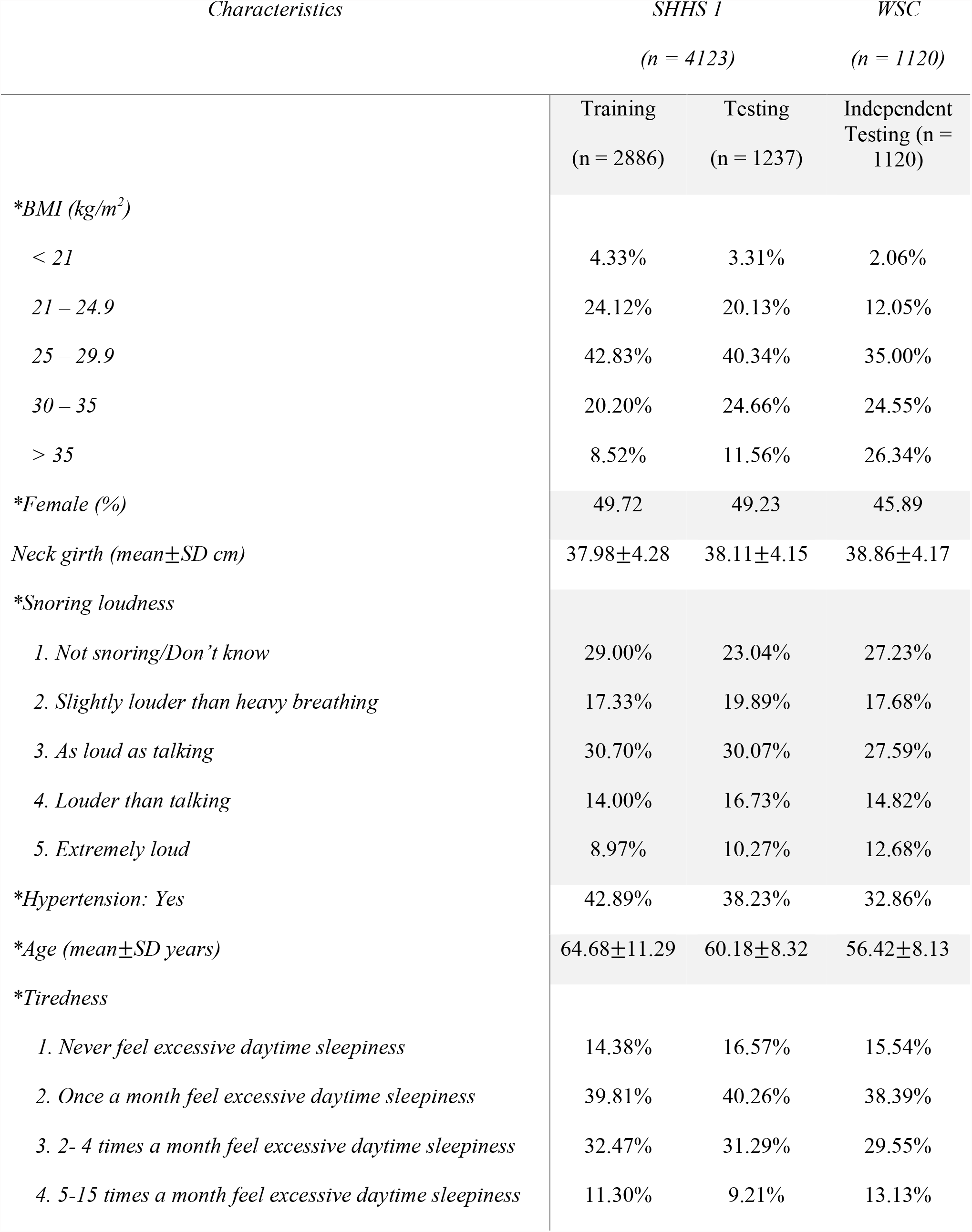
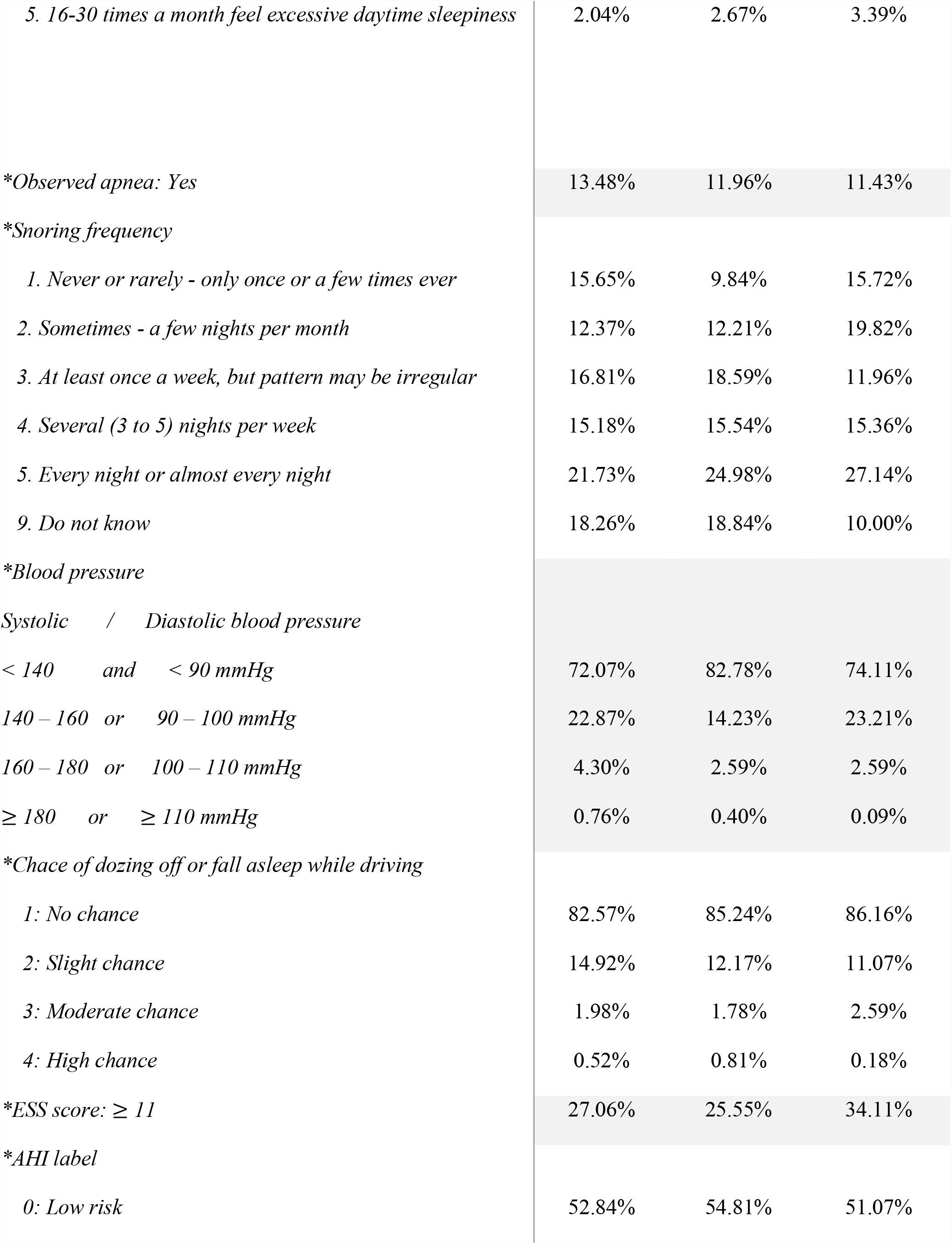
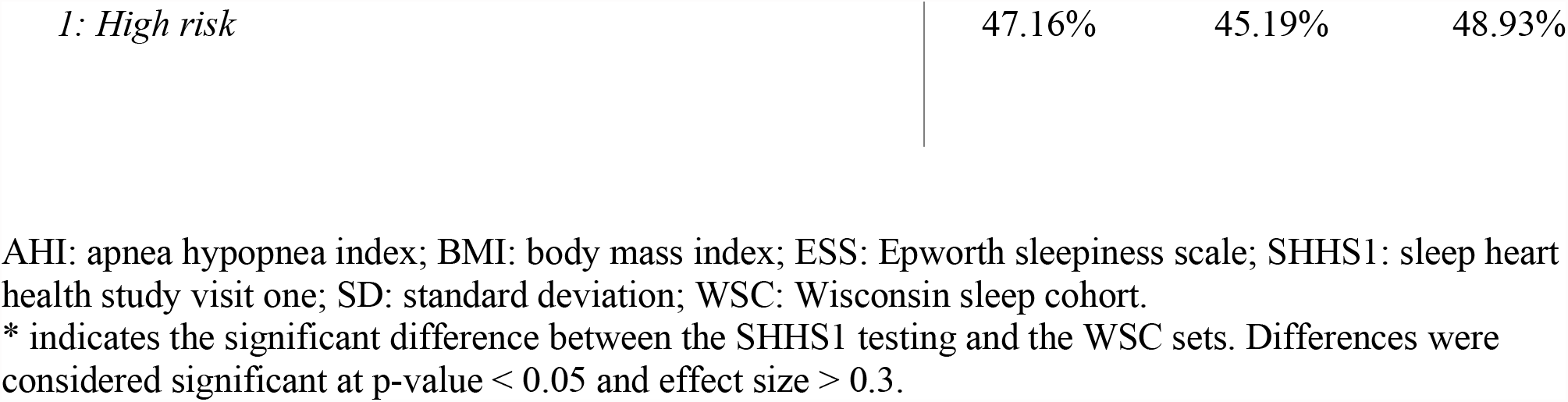
Descriptive characteristics of the datasets

Table 2 shows the importance of risk factors in descending order by MI score. We selected the top 50% (n = 6) features (BMI, gender, neck girth, snoring loudness, hypertension, and age) to develop the machine learning model. The low and high phenotype groups in the SHHS1 training set have different characteristics as shown in Fig. S3.

**Table 2.**
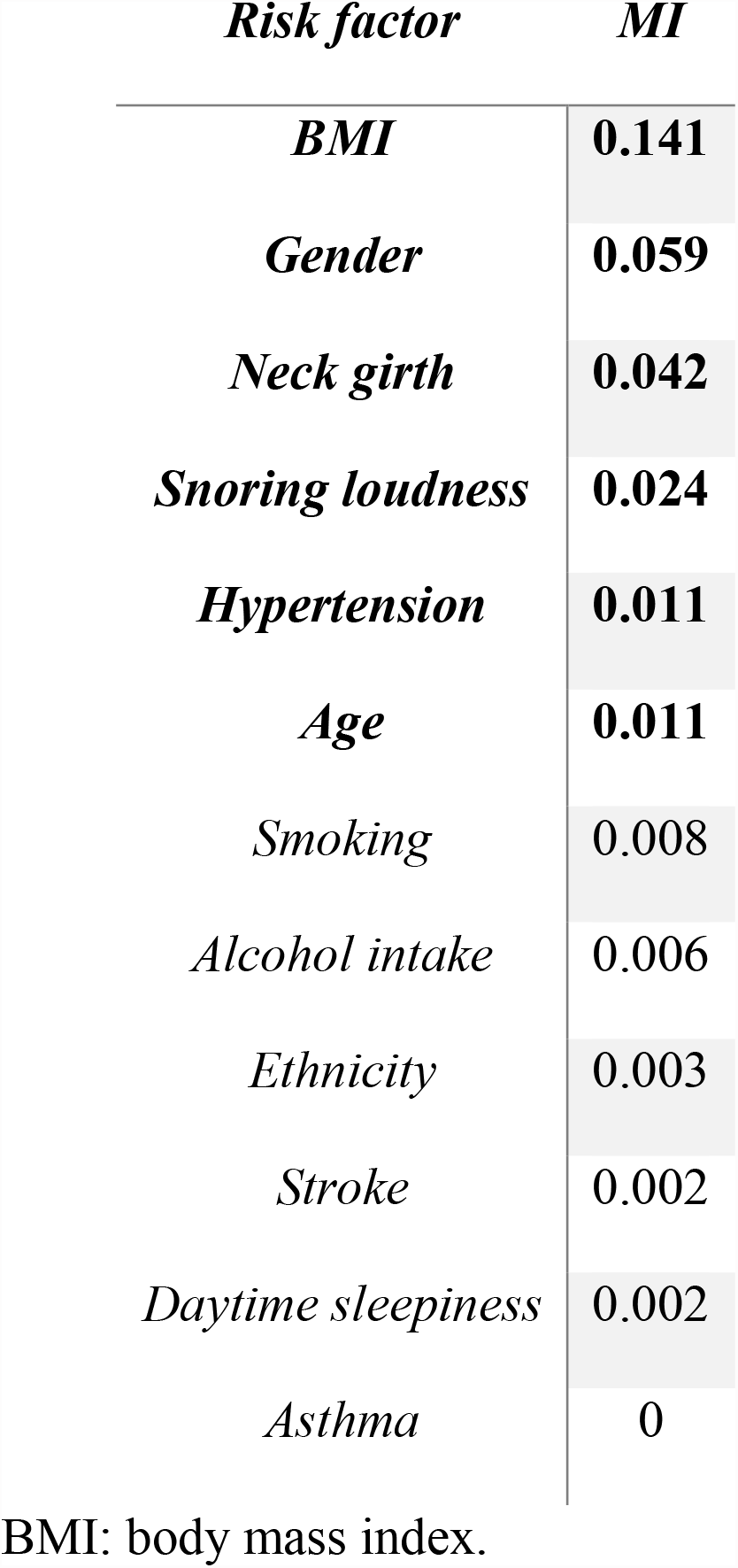
Mutual information score of each risk factor versus apnea-hypopnea index

| *Risk factor* | *MI* |
| --- | --- |
| ***BMI*** | **0.141** |
| ***Gender*** | **0.059** |
| ***Neck girth*** | **0.042** |
| ***Snoring loudness*** | **0.024** |
| ***Hypertension*** | **0.011** |
| ***Age*** | **0.011** |
| *Smoking* | 0.008 |
| *Alcohol intake* | 0.006 |
| *Ethnicity* | 0.003 |
| *Stroke* | 0.002 |
| *Daytime sleepiness* | 0.002 |
| *Asthma* | 0 |

BMI: body mass index.

Table 3 presents the coefficients of two independent logistic regression classifiers to analyze the relationship between the risk factors and the OSA risk. Since the variables were standardized before training, it should be noted that the coefficients shown in Table 3 have been reversed from standardization for interpretation. The logistic regression coefficient showed the expected change in log odds of OSA risk with a risk factor per unit change. Both classifiers had a negative intercept, indicating the odds were against the high OSA risk when values of variables (risk factors) were equal to 0. Hypertension and gender were binary encoded as 0 for non-hypertension and 1 for hypertension, 0 for female and 1 for male, respectively. Hypertension, BMI, age, neck girth and gender demonstrated contributions to the OSA risk due to positive coefficients. The snoring loudness was binary encoded to three variables ranging from 000 to 100 to represent 5 statuses shown in Table S1. Although coefficients of snoring loudness 1 were close to 0 for both groups, the positive weights of snoring loudness 2 and snoring loudness 3 still demonstrated an association between snoring loudness and OSA risk. Both classifiers showed similar weights across the risk factors except for age and snoring loudness 1. The low phenotype group had a coefficient of 0.051 for age while the high phenotype group only had a value of 0.026. The coefficient of snoring loudness 1 in the low phenotype group is positive while it is negative in the high phenotype group, indicating the participants who do not snore may still have high OSA risk in the low phenotype group.

**Table 3.** Coefficients of logistic regression classifiers for high phenotype and low phenotype groups

|  | *low phenotype* | *high phenotype* |
| --- | --- | --- |
| *Intercept* | -9.356 | -6.772 |
| *Hypertension* | 0.226 | 0.169 |
| *BMI* | 0.067 | 0.063 |
| *Age* | 0.051 | 0.026 |
| *Neck girth* | 0.087 | 0.071 |
| *Gender* | 0.673 | 0.596 |
| *Snoring Loudness 1* | 0.121 | -0.143 |
| *Snoring Loudness 2* | 0.609 | 0.495 |
| *Snoring Loudness 3* | 0.291 | 0.371 |

BMI: body mass index; The snoring loudness was binary encoded to three variables ranging from 000 to 100 to represent 5 statuses shown as Table S1.

The AUROC of the BASH-GN and other 4 questionnaires on SHHS1 and WSC testing sets are shown in Fig. 2 (a) and (b), respectively. Table 4 shows the AUROC and AUPRC of the BASH-GN and other 4 questionnaires. The optimal threshold shown in Fig. 2 (a) and (b) was chosen according to the geometric mean for the balance of sensitivity and specificity, which was calculated by the maximum values of true positive rate * (1 – false positive rate). With a selected threshold = 0.427, our model reached a sensitivity of 0.77 and a specificity of 0.68 on the SHHS 1 testing set and had a 0.69 sensitivity and a 0.72 specificity on the WSC testing set. The BASH-GN model had consistently better performance in terms of AUROC on both testing sets. Compared to the other comparison questionnaires, the BASH-GN model demonstrated better performance in terms of the AUROC and AUPRC on both testing sets. The result also indicated a stable performance of the BASH-GN model between two testing sets on AUROC (SHHS1: 0.78, WSC: 0.76) and AUPRC (SHHS1: 0.72, WSC:0.74), whereas the performance of comparison questionnaires fluctuated when the data label distribution varied.

**Fig. 2.**
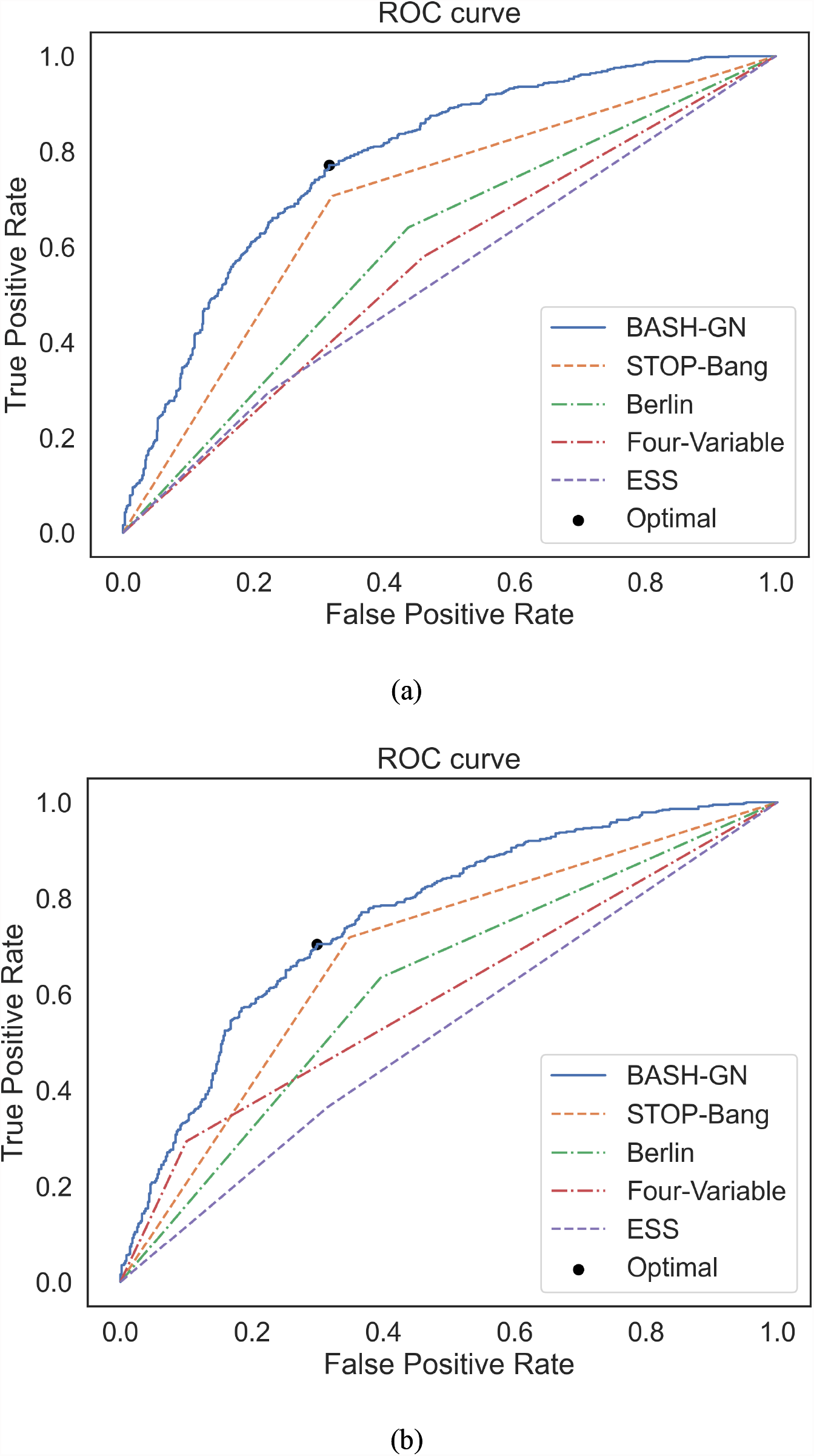
The receiver operation characteristics (ROC) curve for OSA risk classification. (a) SHHS 1 testing set. (b) WSC testing set.

**Table 4.**
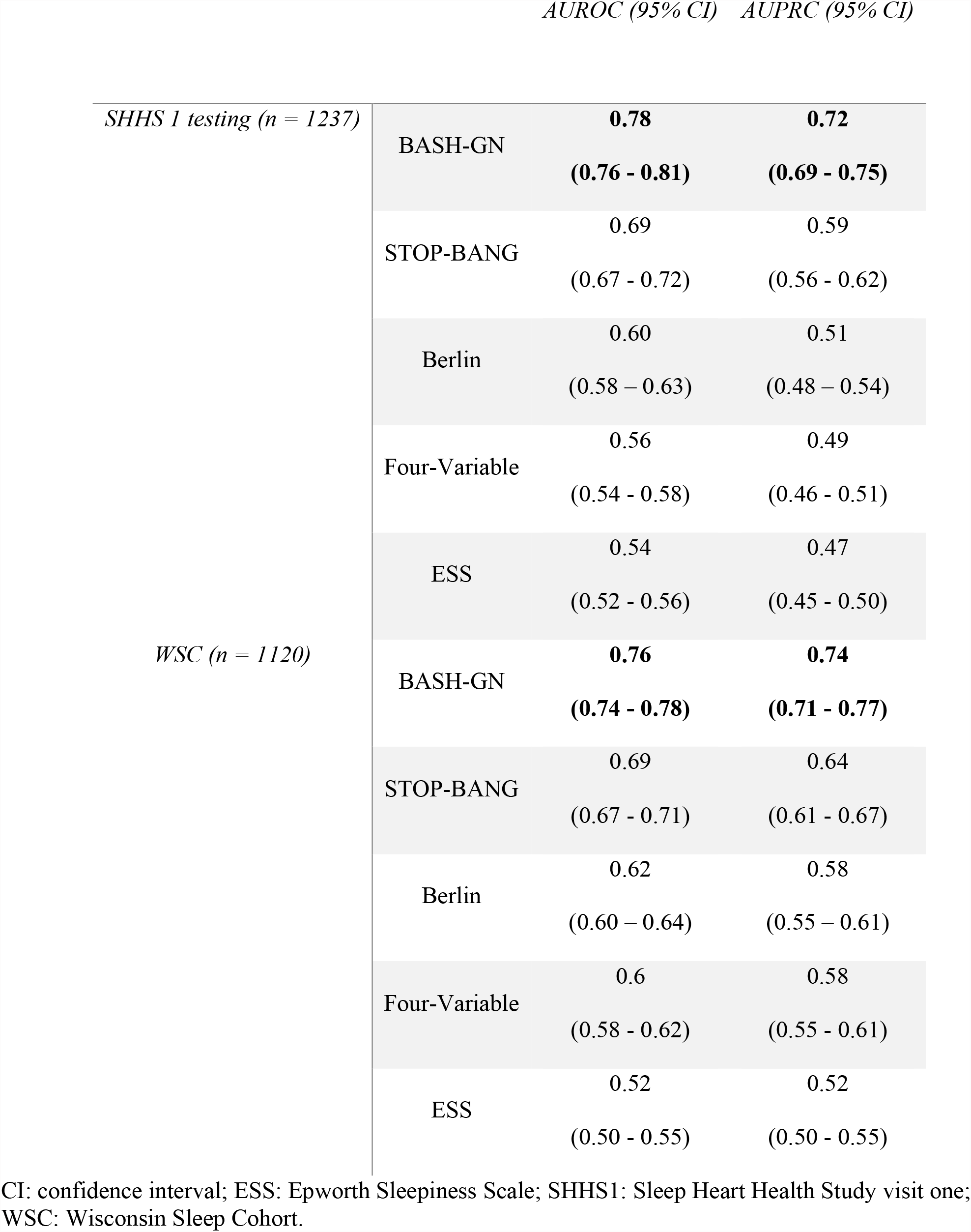
Performances of BASH-GN model and other questionnaires on mean area under the receiver operating characteristics (AUROC) and area under the precision-recall curve (AUPRC)

## Discussion

In this study, we developed the BASH-GN, a 6-item questionnaire, to predict moderate to severe OSA risk by considering risk factor subtypes based on a machine learning model. According to the symptoms of participants, the model classified the subjects into two different groups, a low phenotype and a high phenotype, followed by two independent logistic regression classifiers for binary OSA risk prediction. The model was trained on a subset of the SHHS 1 (n = 2886) dataset, with a balanced distribution of binary OSA labels, and obtained a 0.78 (95% CI: 0.76-0.81) AUROC, and a 0.72 (95% CI: 0.69-0.75) AUPRC on the holdout testing set (n = 1237). We also evaluated the generalizability of the model on the independent WSC dataset (n = 1120). The model demonstrated a similar performance with an AUROC of 0.76 (95% CI: 0.74-0.78) and an AUPRC of 0.74 (95% CI: 0.71-0.77). This study demonstrated that the BASH-GN had a consistent and better performance on both testing sets regarding the AUROC and AUPRC compared to alternative questionnaires.

The proposed BASH-GN is simpler and easier to gather the data compared to alternative questionnaires. The Four-Variable only has 4 items, but it may be less useful in as much as systolic and diastolic blood pressures are required for assessment. Both ESS and STOP-BANG questionnaires require participants to answer 8 questions, while the Berlin may need up to 10 items. Moreover, STOP-BANG and Berlin also require information on observed stop breathing. However, Nagappa et al. has noted that observed stop breathing may not be accurately captured in the absence of participants’ bed partners [29]. In contrast, the variables in BASH-GN are easier to assess.

We found that the intercept of the low phenotype group is lower than that of the high phenotype group. The low phenotype group had an intercept of -9.356, while the high phenotype group had an intercept of - 6.772. Except the snoring loudness, the rest of the coefficients of the low phenotype group are higher than that of the high phenotype group. For example, the coefficient of age for the low phenotype group was 0.051 whereas it was 0.026 for the high phenotype group. Furthermore, we found the coefficient of snoring loudness 1 (Don’t know/Not snoring) of the high phenotype group was -0.143, indicating a decreased odds of OSA for participants without snoring. In contrast, the coefficient of snoring loudness 1 was positive in the low phenotype group, implying that many participants with high OSA risk in the low phenotype group may not snore. Therefore, taking OSA subtypes into account to identify OSA risk is important.

We have demonstrated that the BASH-GN questionnaire which uses a machine learning derived algorithm is more accurate in predicting the presence of moderate to severe OSA. It is currently available on the web at: https://c2ship.org/bash-gn, and could easily be incorporated into an app for use on mobile devices. Therefore, it could be conveniently accessed by primary care practitioners and other clinicians for office screening as part of routine office visits. Furthermore, electronic medical records (EMR) are now incorporating practice messages whereby “flags” appear when a patient’s medical record is opened to remind clinicians to address an important health care issue. The BASH-GN could be likewise incorporated into the EMR as a means of increasing the recognition and eventual treatment of OSA.

Several limitations of our study should be noted. First, the BASH-GN model was trained for OSA risk prediction only. Further verifications may be needed for other types of sleep-disordered breathing classification. Second, it is known the severity of OSA is classified as none, mild, moderate, and severe. We only tested the binary prediction with a cut-off value of 15 for AHI which may be less informative for screening. However, this may not be clinically important because the need to treat less severe OSA is still unclear [30]. Importantly, the model was tested developed and tested on 2 general population datasets. Further testing on clinical populations is needed.

In conclusion, the BASH-GN questionnaire which incorporates OSA subtype information improves the accuracy of OSA screening compared to other commonly used screening instruments. It has the potential to be an important clinical tool in the identification of patients with OSA.

## Data Availability

All data are available online at https://sleepdata.org

## Declarations

### Ethical approval

For this type of study formal consent is not required.

### Informed consent

Informed consent was obtained from all individual participants included in the study.

### Data availability statements

The datasets analyzed during the current study are publicly accessible via https://sleepdata.org/datasets/shhs and https://sleepdata.org/datasets/wsc.

## Supplementary

### Variable Description

**Table S1:**
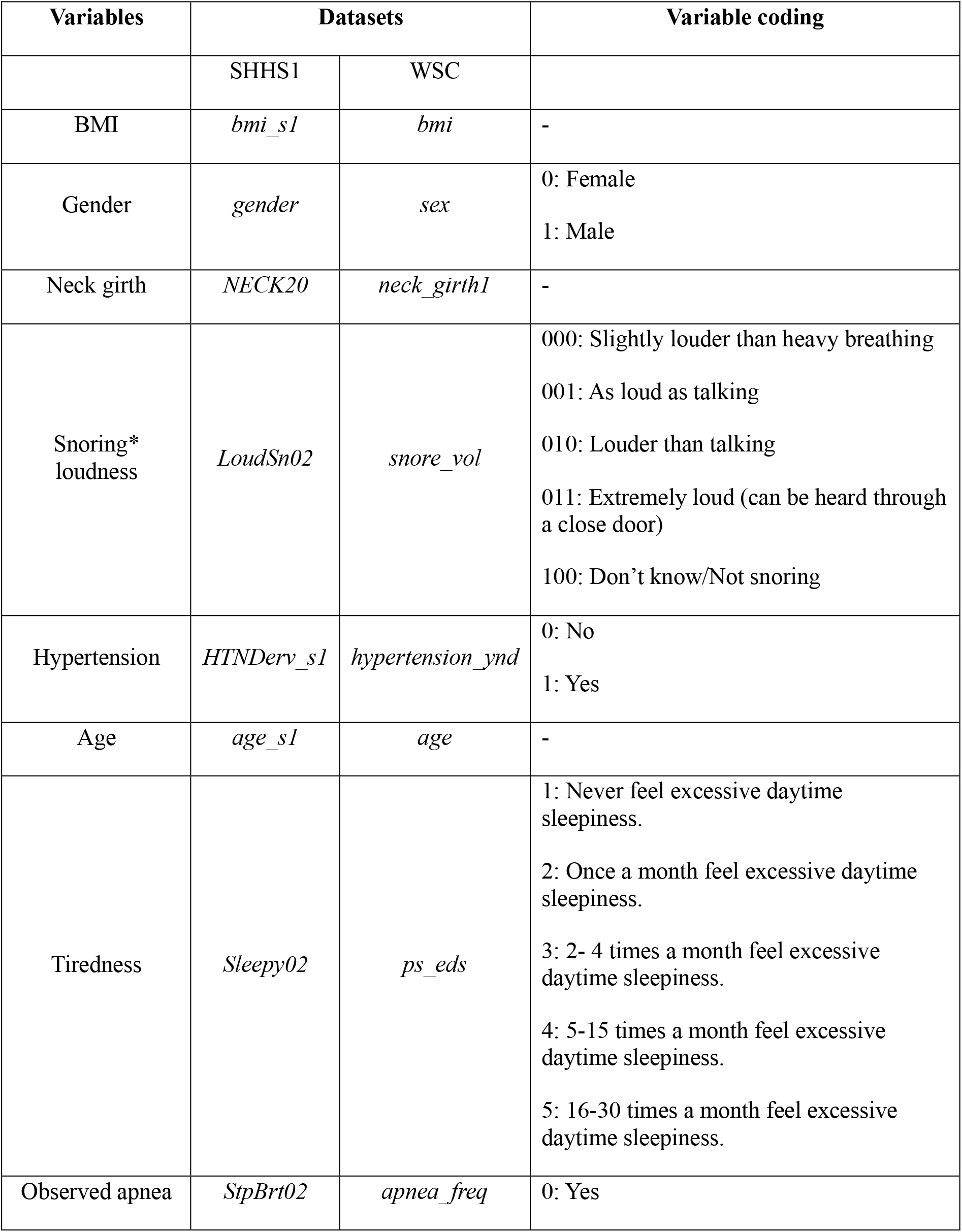
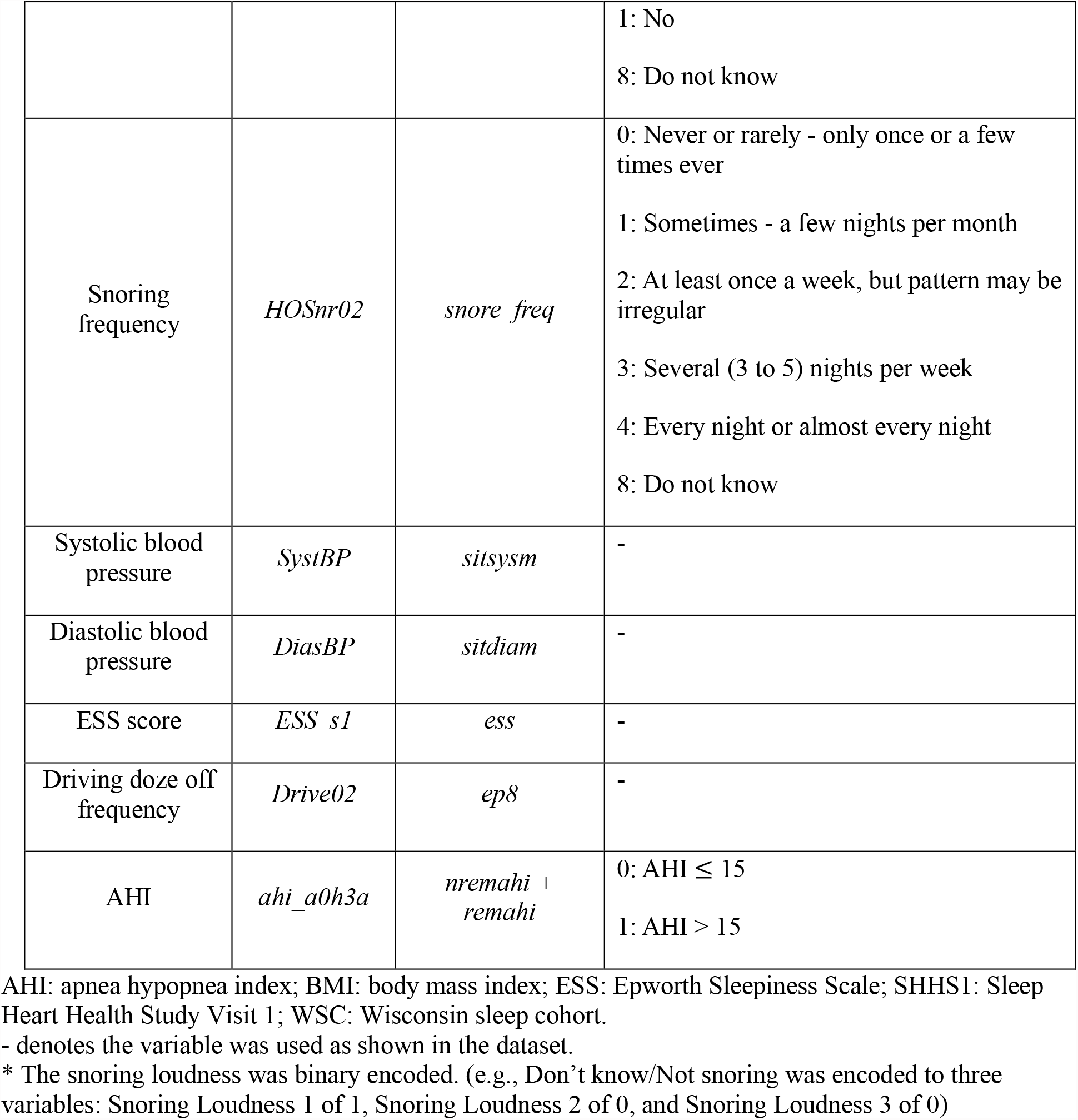
Variables used in both datasets

### Questionnaire

The questionnaire consisted of six questions using selected OSA risk factors. We used the same questions/options described by SHHS 1 dataset except for the snoring loudness. In the SHHS sleep habits questionnaire, the answer to snoring loudness will be blank if participants selected “Not snoring” in the previous snoring frequency question. Therefore, we added “Not snoring” and combined it with “Don’t know” as one option for snoring loudness. Questions are listed as shown in Table S2.

**Table S2.** Proposed questionnaire preview

| Item | Question | Choices/Answer |
| --- | --- | --- |
| 1 | Age |  |
| 2 | Gender | 1. Female 2. Male |
| 3 | Neck circumference in cm |  |
| 4 | Body Mass Index |  |
| 5 | Do you have high blood pressure or being treated with Hypertension medicines? | 1. No 2. Yes |
| 6 | How loud is your snoring? | 1. Slightly louder than heavy breathing 2. As loud as talking<br>3. Louder than talking 4. Extremely loud (can be heard through a close door) 5. Don't know/Not snoring |

### Phenotype group threshold selection

The SHHS 1 training set (n = 2875) was used to decide the threshold of phenotype group categorization. Each question in Table S2 was assigned a score of 1 and the following cutoffs were used for scoring: age > 50, gender = male, neck circumference > 40 cm, body mass index > 35, high blood pressure = Yes, snoring is louder than talking. Categorization performance was measured by the area under the receiver operating characteristics curve for threshold selection, as shown in Fig. S1. The optimal threshold (= 3) was selected based on the maximum product of true positive rate and (1 - false positive rate) among all threshold settings.

**Fig S1:**
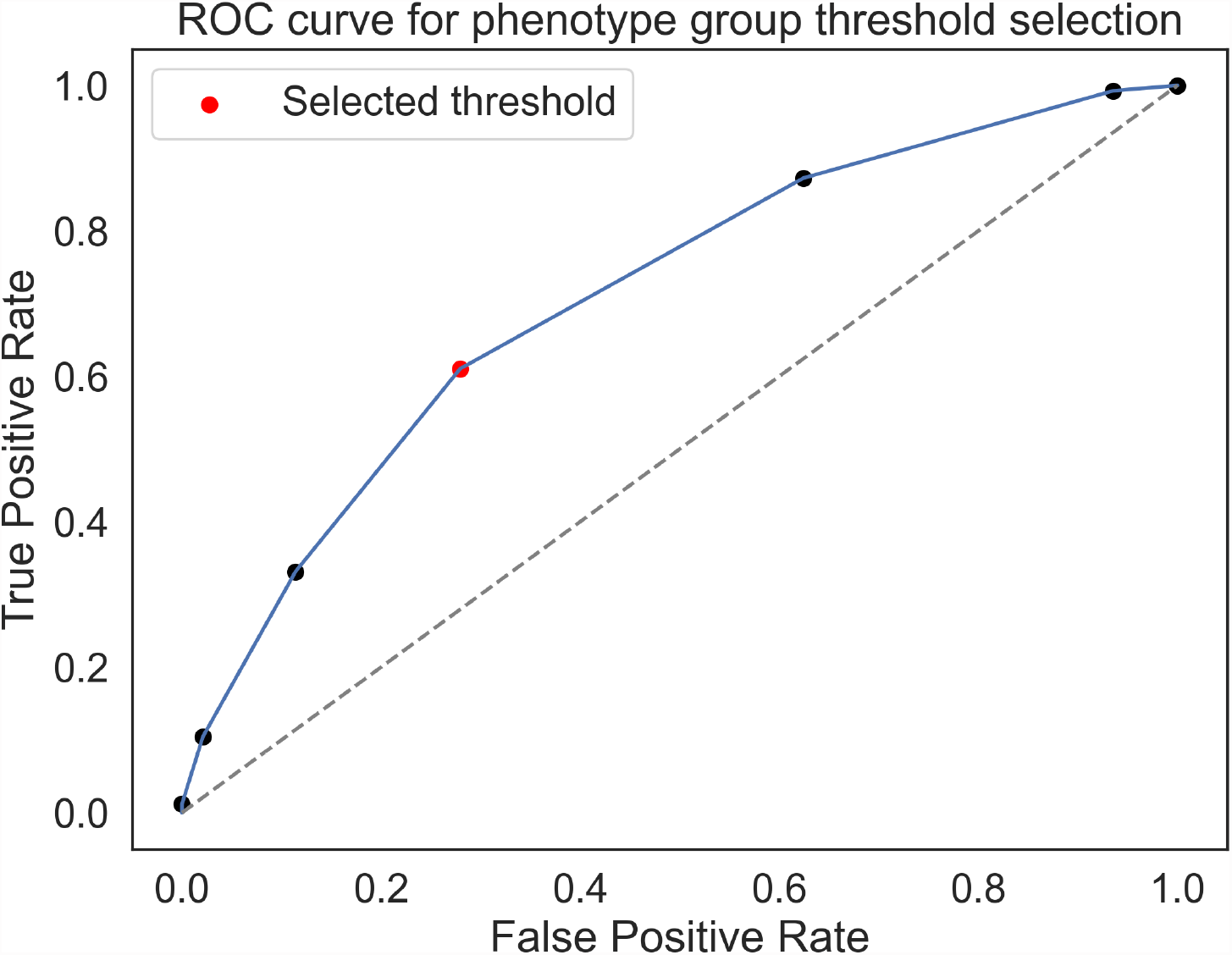
The receiver operation characteristics (ROC) curve for phenotype group threshold selection. The selected threshold (= 3) was marked as red.

### Characteristics comparison between two phenotype groups

The z-scores were calculated via the following equation:

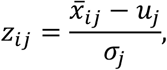

where i = 1, 2 represents the low and the high phenotype group, respectively; j denotes j^th^ risk factor; *z*_*ij*_represents the j^th^ risk factor’s z-score of i^th^ phenotype group; *u*_*j*_ and *σ*_*j*_ denote the mean and standard deviation of j^th^ risk factor in SHHS 1 training set, respectively; 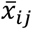 is mean of j^th^ risk factor of i^th^ phenotype group. The z-score represents the relative change of mean between low and high phenotype groups. The positive value of a z-score reflects an increase from the mean value, whereas the negative values of a z-score reflect a decrease from the mean score of the training set. The low phenotype and high phenotype groups have different characteristics. As shown in Figure S2, the high phenotype group had higher z-scores across all the selected risk factors. In contrast, the means of the low phenotype group were lower than that of the overall training set, which can be challenging to identify using the same standard for classification.

**Fig. S2:**
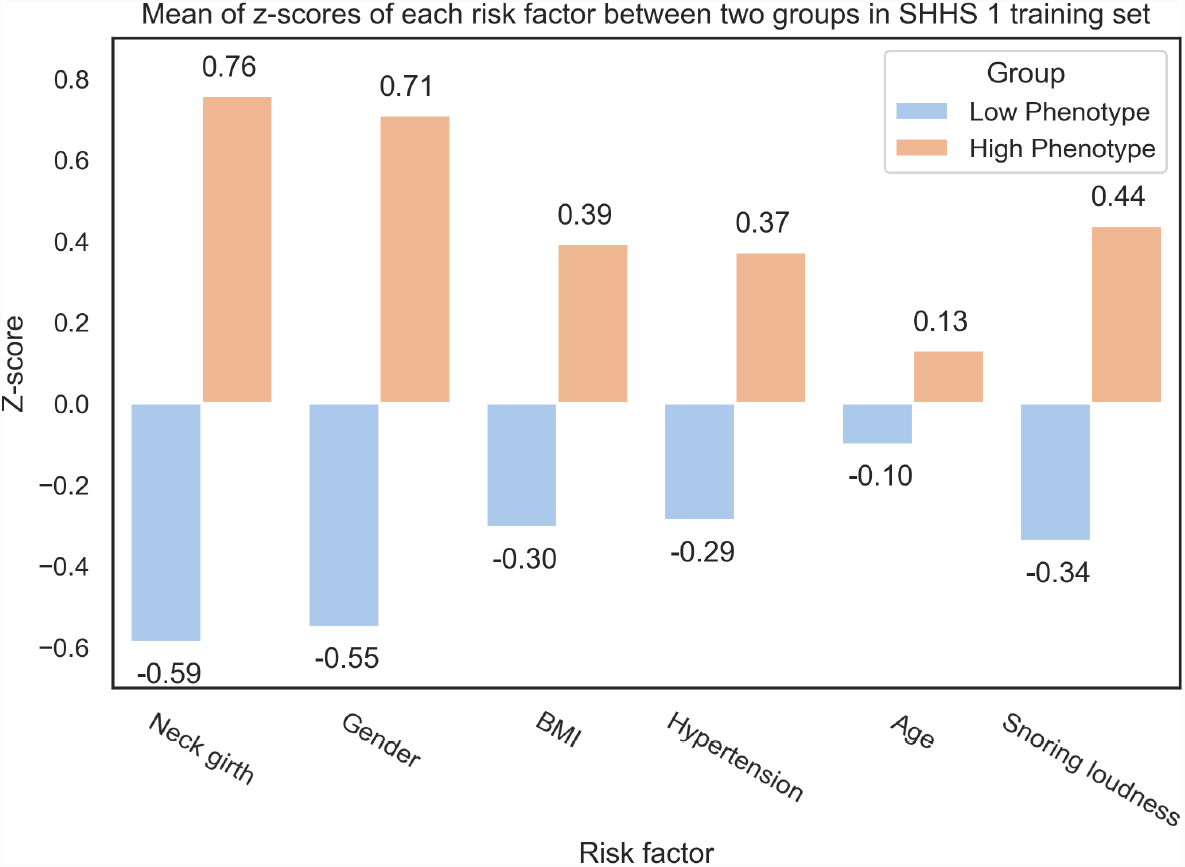
Z-scores of risk factors between two groups in SHHS 1 training set

### 10-fold cross validation result for algorithms selection

We used stratified 10-fold cross validation to select the algorithms with the best AUROC mean for two phenotype groups. Fig. S3 shows that the logistic regression (LR) had the best performance regarding AUROC mean in both phenotype groups.

**Fig. S3.**
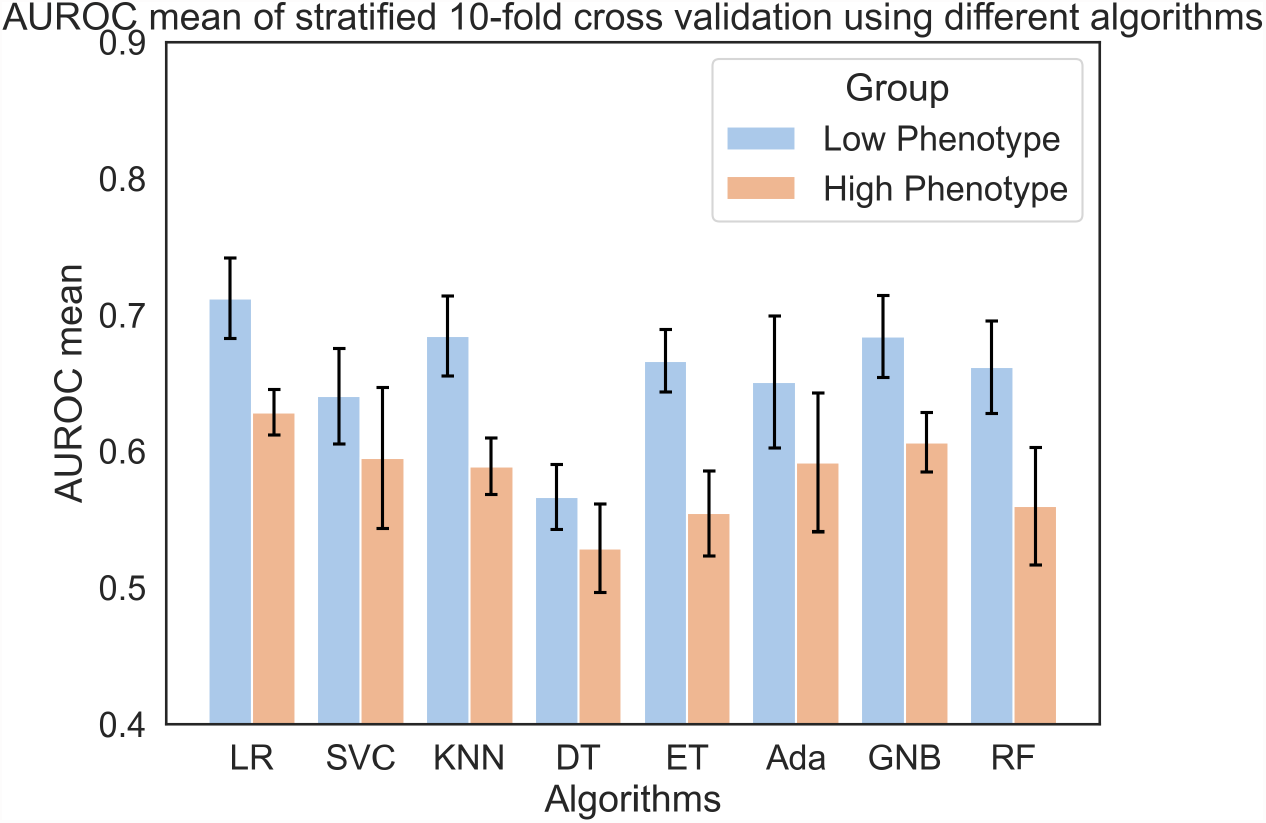
Area under the receiver operating characteristic (AUROC) mean of stratified 10-fold cross validation using different algorithms. Error bar indicates the standard deviation (SD). (Expressed as algorithm (G1 = Symptomatic: mean ± SD, G2 = Minimal symptomatic: mean ± SD)). LR = logistic regression; SVC = support vector classifier; KNN = K-nearest neighbors; DT = decision tree; ET = extra tree; AB = Ada boost; GNB = Gaussian Naïve Bayes; RF = random forest

### Coding of the STOP-BANG, ESS, Berlin, and Four-variable questionnaires

To compare with the STOP-BANG questionnaire, the risk of OSA was calculated based on 8 questions. Specifically, 1) Snore was defined as yes if snoring loudness is louder than talking; 2) Tiredness was defined as positive if participants reported excessively sleepy during the day more than 5 times/month; 3) Observed stop breathing was considered affirmative if participants answered *yes* to “Are there times when you stop breathing during your sleep?” in SHHS1 or if participants denoted stop breathing experience in “According to what others have told you, or to your own awareness, how often, if ever, do you have momentary periods during sleep when you stop breathing or you breathe abnormally?” in WSC; 4) Hypertension was considered affirmative if participants indicated hypertension history or being treated with hypertension medicine; 5) BMI ≥35 kg/m^2^ was defined as positive; 6) Age ≥50 was considered as an affirmative answer; 7) Neck girth was considered affirmative if it is over 40 cm; 8) Male was defined as positive for gender. Low risk of OSA was defined as less than 3 affirmative answers while the participant will be considered as high risk of OSA if there were more than 2 affirmative answers.

The ESS was completed by the SHHS and WSC participants and the total ESS score ranges from 0 – 24. We applied the threshold of 11[26] to classify the subjects into low risk or high risk of OSA.

The Four-variable questionnaire was assessed through BMI, blood pressures, gender, and snoring frequencies. Each variable was initially classified into different categories with assigned scores; then a linear equation was utilized to calculate the total score of the subjects. Specifically, 1) BMI was assigned a value from 1 to 6 for 6 ranges (<21, 21-23, 23-25, 25-27, 27-29, ≥30 kg/m^2^), respectively; 2) according to systolic and diastolic blood pressure, blood pressures were defined as 4 intervals (systolic < 140 or diastolic < 90, systolic 140-160 or diastolic 90-100, systolic 160-180 or diastolic 100-110, systolic ≥180 or diastolic ≥110) and assigned a score of 1 to 4 for each category; 3) gender was assigned a score of 0 for females and 1 for males; 4) snoring frequency was assigned a score of 0 if the participants snored less than 3 nights per week, and a score of 1 for participants who snored ≥3 nights per week. Finally, we used the equation, BMI score + blood pressure score + 4 * (gender score) + 4 * (snoring frequency score), to get the total score, and subjects were divided into low risk and high risk of OSA using a threshold of 14.

Berlin questionnaire (BQ) included 10 questions in three sections related to the snoring, daytime fatigue, and obesity or hypertension. Each section was evaluated separately. If two sections were assessed as positive, the subject was classified as high risk of OSA. For the snoring section, “*Do you snore?*” was considered as *Yes* and assigned 1 score if participants denoted a snoring loudness of “How loud is your snoring” in both datasets. As for question 2, “*Your snoring is*,” would be assigned 1 score if snoring loudness was louder than talking. “*How often do you snore?*” would be considered affirmative and add 1 score if the snoring frequency was higher than 3 times per week. “*Has your snoring ever bothered other people?*” would assign 1 score if snoring loudness was louder than talking. There was no variable about observed stop breathing frequency in SHHS1 and WSC that can match the question “*Has anyone noticed that you stop breathing during your sleep?*” in BQ. Thus, if participant was observed stop breathing, this question was assigned a score of 2 according to the instruction of BQ. The snoring category was considered positive if total assigned score was higher than 2. For the daytime fatigue category, due to the similarity, the question “*How often do you feel tired or fatigued after your sleep?*” and “*During your waking time, do you feel tired, fatigued or not up to par?*” were combined and assigned a score of 2 if participant reported excessively sleepy during the day more than 16 times/month. “*Have you ever nodded off or fallen asleep while driving a vehicle*” was assigned 1 score if driving doze off frequency was equal to or higher than “Slight Chance”. The daytime fatigue category was considered as positive if the assigned score is 2 or more in this section. Lastly, if participants indicated hypertension history or being treated with hypertension medicine, or BMI was higher than 30 kg/m^2^, category 3 was considered positive.

